# Spatial immunosampling of MRI-defined glioblastoma regions reveals immunologic fingerprint of non-contrast enhancing, infiltrative tumor margins

**DOI:** 10.1101/2023.03.09.23285970

**Authors:** Matthew M. Grabowski, Dionysios C. Watson, Kunho Chung, Juyeun Lee, Defne Bayik, Adam Lauko, Tyler Alban, Jan Joseph Melenhorst, Timothy Chan, Justin D. Lathia, Manmeet S. Ahluwalia, Alireza M. Mohammadi

**Affiliations:** Cleveland Clinic Lerner Research Institute, Cleveland, OH, USA; Case Comprehensive Cancer Center, Cleveland, OH, USA; Rose Ella Burkhardt Brain Tumor and Neuro-Oncology Center, Department of Neurosurgery, Cleveland, OH, USA; Case Western Reserve University, Cleveland, OH, USA; University Hospitals Cleveland Medical Center, Cleveland, OH, USA; Miami Cancer Institute, Baptist Health South Florida, Miami, FL, USA; Medical Scientist Training Program, Case Western Reserve University School of Medicine, Cleveland, Ohio, USA

**Keywords:** tumor microenvironment, glioblastoma, spatial heterogeneity, tumor-infiltrating immune cells, image-guided biopsy

## Abstract

Glioblastoma (GBM) treatment includes maximal safe resection of the core and MRI contrast-enhancing (CE) tumor. Complete resection of the infiltrative non-contrast-enhancing (NCE) tumor rim is rarely achieved. We established a safe, semi-automated workflow for spatially-registered sampling of MRI-defined GBM regions in 19 patients with downstream analysis and biobanking, enabling studies of NCE, wherefrom recurrence/progression typically occurs. Immunophenotyping revealed underrepresentation of myeloid cell subsets and CD8+ T cells in the NCE. While NCE T cells phenotypically and functionally resembled those in matching CE tumor, subsets of activated (CD69^hi^) effector memory CD8+ T cells were overrepresented. Contrarily, CD25^hi^ Tregs and other subsets were underrepresented. Overall, our study demonstrated that MRI-guided, spatially-registered, intraoperative immunosampling is feasible as part of routine GBM surgery. Further elucidation of the shared and spatially distinct microenvironmental biology of GBM will enable development of therapeutic approaches targeting the NCE infiltrative tumor to decrease GBM recurrence.

## INTRODUCTION

Glioblastoma (GBM) is the most common primary malignant brain tumor in adults^1^, with a near 100% recurrence rate after standard care with surgery, radiation, and chemotherapy^2^. While immune checkpoint inhibitors (e.g. anti-PD-1, -PD-L1, -CTLA-4) have improved outcomes in numerous other cancer types, they have shown limited efficacy in GBM^3^. Implementing checkpoint inhibition and other forms of immunotherapy in GBM has been challenging due to the presence of a highly immunosuppressive tumor microenvironment, driven by tumor cells and numerous immunosuppressive mechanisms in the brain^3,4^. An additional layer of complexity arises from spatial heterogeneity of the immune microenvironment, resulting from physical microenvironmental factors (such as hypoxia, the blood brain barrier, etc.), tumor cell heterogeneity, and varying interactions with brain-resident cells^4^. Further complicating the rational design of GBM immunotherapy are technical factors associated with tissue collection and handling that can harm cells in the tissue sample (including immune cells) leading to analytical bias with commonly utilized techniques of flow cytometry and single cell sequencing^5^. Accurately elucidating immune microenvironment spatial heterogeneity in clinically meaningful contexts has the potential to inform future GBM trials, especially those utilizing immunotherapy.

In the context of surgical resection—the first step in standard GBM management—tumors are segmented based on contrast-enhanced MRI, with the tumor core and contrast-enhancing (CE) regions being targeted for complete resection^2^. However, previous studies have shown that maximal safe resection of adjacent T2/FLAIR hyperintense, non-contrast-enhancing (NCE) tumor regions further improves patient outcomes by targeting the leading edge of the infiltrative tumor, where the majority of recurrences occur^6,7^. In many cases though, the extent of resection is limited by eloquent regions of brain and the need to preserve neurological function, resulting in earlier progression which is only partially mitigated by adjuvant chemoradiotherapy^6,7^. While previous studies have described spatial heterogeneity by various segmentation strategies, how this relates to the clinically relevant MRI-defined regions remain poorly understood.

In this prospective study, we developed a streamlined workflow for MRI-guided, spatially-registered intraoperative sampling of the core, CE, and NCE regions for downstream immune profiling and biobanking. Given the established link between CE and inflammation in MRI^8^, we hypothesized that the NCE would be characterized by a pauci-immune microenvironment, with T lymphocytes lacking activation/exhaustion markers that are hallmarks of tumor-infiltrating lymphocytes. While we did observe a lower abundance of leukocytes in the NCE, they phenotypically and functionally resembled immune cell populations found in the core and CE regions. On the other hand, specific T cell subsets displayed differential abundance in the NCE, including activated effector memory CD8^+^ cells. These data suggest that designing adjuvant immunotherapeutic strategies that enable this region’s immune cells to target residual GBM in the NCE could be a potentially promising approach to limit recurrence of this deadly disease.

## RESULTS

### Optimized workflow facilitates high-viability, spatially-registered GBM sampling

19 patients were included in this study (**Table S1**), and one patient was excluded from analysis due to postoperative diagnosis of IDH-mutant tumor. Tissue acquisition was surgically feasible at all annotated regions using the NICO Myriad device (**Figure 1A-B**) and on average, sampling added 10 minutes to surgical time. No adverse events attributable to the device or acquisition protocol were observed. The workflow we developed obtained sufficient cells for immediate flow cytometry in 96.5% (55/57) samples. 86% (49/57) of samples were sufficient to additionally biobank viably frozen single-cell suspensions, while 98.2% of samples were sufficient to generate flash frozen tumor specimens. We next assessed the viability of fresh single-cell suspensions by flow cytometry using a viability dye as a quality metric of the GBM sampling workflow we developed. Median viability was >80% in all sampled regions, with the NCE displaying the highest viability (median 98%, 95% confidence interval [CI] 91-99%; **Figure 1C**). Most of the very low viability samples (<50%) were found in the core (4 of 6), possibly reflecting sampling in or around necrotic regions characteristic of GBM rather than a result of sample processing.

**Figure 1:**
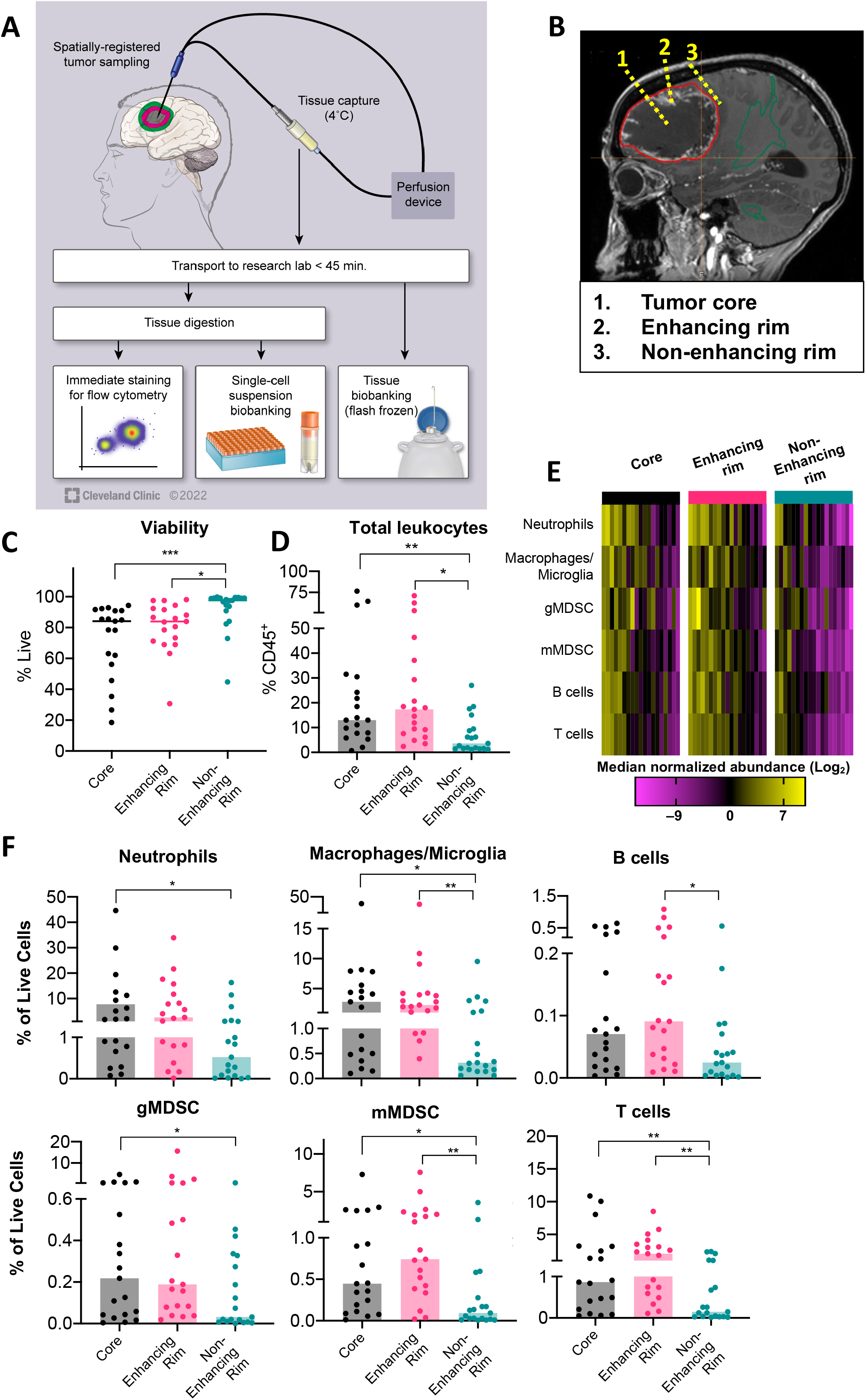
Spatially-registered GBM sampling workflow enables studies of regional GBM heterogeneity. (**A**) Schematic overview of workflow. (**B**) Sagittal MRI plane depicting 3 NICO Myriad trajectories (yellow dotted lines) to obtain spatially-registered samples from human GBM (red outline). 1=core, 2=CE, 3=NCE. (**C**) Viability of fresh single-cell suspensions analyzed by flow cytometry, as a percent of total singlet cell events. (**D**) Abundance of total CD45-expressing leukocytes in each tumor region, as a percent of live cells. (**E**) Overview of spatial heterogeneity in leukocyte abundance (as percent of total live cells), normalized to median of each cell type. (**F**) Abundance of individual immune cell subsets across tumor regions, as a percent of live cells. *p <0.05, **p<0.01, ***p<0.001; Freidman test.

### NCE tumor has lower frequency of infiltrating immune cells

Immediate immunophenotyping of single-cell suspensions allowed us to assess the immune microenvironment of the three designated tumor regions (see **Supplementary Figure S1** for gating strategy), including cells prone to loss during freeze/thaw of banked samples. The proportion of live cells that were leukocytes varied greatly, from 0.68 – 76.5% (**Figure 1D**). In line with our originating hypothesis, the overall abundance of tumor-infiltrating immune cells was found to be decreased in the NCE region (median 3.6%) compared to the CE region (median 17.3%) and core (median 13%) (**Figure 1D-E**). Neutrophils, macrophages/microglia, granulocytic and monocytic myeloid-derived suppressor cells (g [or PMN] and mMDSC), and T cells were all underrepresented in the NCE (**Figure 1F**).

### T cells in NCE tumor retain characteristics of CE regions

Given the observed decrease in the abundance of multiple immune cell subset in the NCE, we hypothesized that this region would be immunologically “cold” and characterized by T cells with lower levels of activation and exhaustion as compared to the main tumor mass (CE and core). To test this hypothesis, we performed in-depth T cell immunophenotyping. In the initial analysis of freshly isolated cells, we observed a decrease in the CD8^+^:CD4^+^ T cell ratio in the NCE region (**Figure 2A**). In CD4^+^ T cells, an increased relative frequency of effector memory (CD45RA^lo/-^CCR7^lo/-^) cells was observed at the expense of central memory (CD45RA^lo/-^CCR7^+^) cells in the NCE (**Figure 2B**). While the same pattern was observed among CD8+ T cells, this did not reach statistical significance (**Figure 2C**). In a subset of patients, we further assessed a more expanded panel of activation and exhaustion markers in T cells from biobanked (viably frozen) samples (see gating strategy in **Supplementary Figure S2**) and found that an increased relative frequency of PD-1^+^TOX^+^ cells of CD4^+^ and CD8^+^ T cells in the CE compared to the core (**Figure 3A-B**). Furthermore, we observed increase in relative frequency of PD-1^+^CD38^+^ cells in CD8^+^ T cells in the CE and NCE compared to the core (**Figure 3B**). However, we found that T cells in the NCE phenotypically overall resembled those found in the CE and core when analyzed by manual gating (**Figure 3**). Next, we analyzed the abundance of immune cell subsets in the core, CE and NCE by unsupervised analysis to uncover differences that were not detected by manual gating (**Figure 4** and **Supplementary Figure S3**). From 20 computed clusters by FlowSOM algorithm (**Supplementary Figure S3**), T cells from NCE showed some distinct features, including increased abundance of CD69^hi^PD-1^hi^ or CD69^hi^2B4^hi^ CD8 “Activated” effector memory (CCR7^-^CD45RA^-^) T cells, and TCF1^hi^ CD8 “Naïve-like” central memory (CCR7^+^CD45RA^-^) T cells (**Figure 4A-B**). Contrarily, Ki67^hi^ CD4 “Proliferating” regulatory T cells (CD25^+^Foxp3^+^) showed decreased abundance in NCE compared to the core and CE. (**Figure 4B**). Furthermore, T cells from CE showed increased abundance of Ki67^hi^PD-1^hi^ CD4 “Proliferating” central memory (CCR7^+^CD45RA^-^) and CD69^hi^PD-1^hi^ “Activated” CD4 central memory (CCR7^+^CD45RA^-^) T cells (**Figure 4B**), but TCF1^hi^ “Naïve-like” CD4 central memory (CCR7^+^CD45RA^-^) T cells showed decreased abundance in CE compared to the core and NCE (**Figure 4B**).

**Figure 2:**
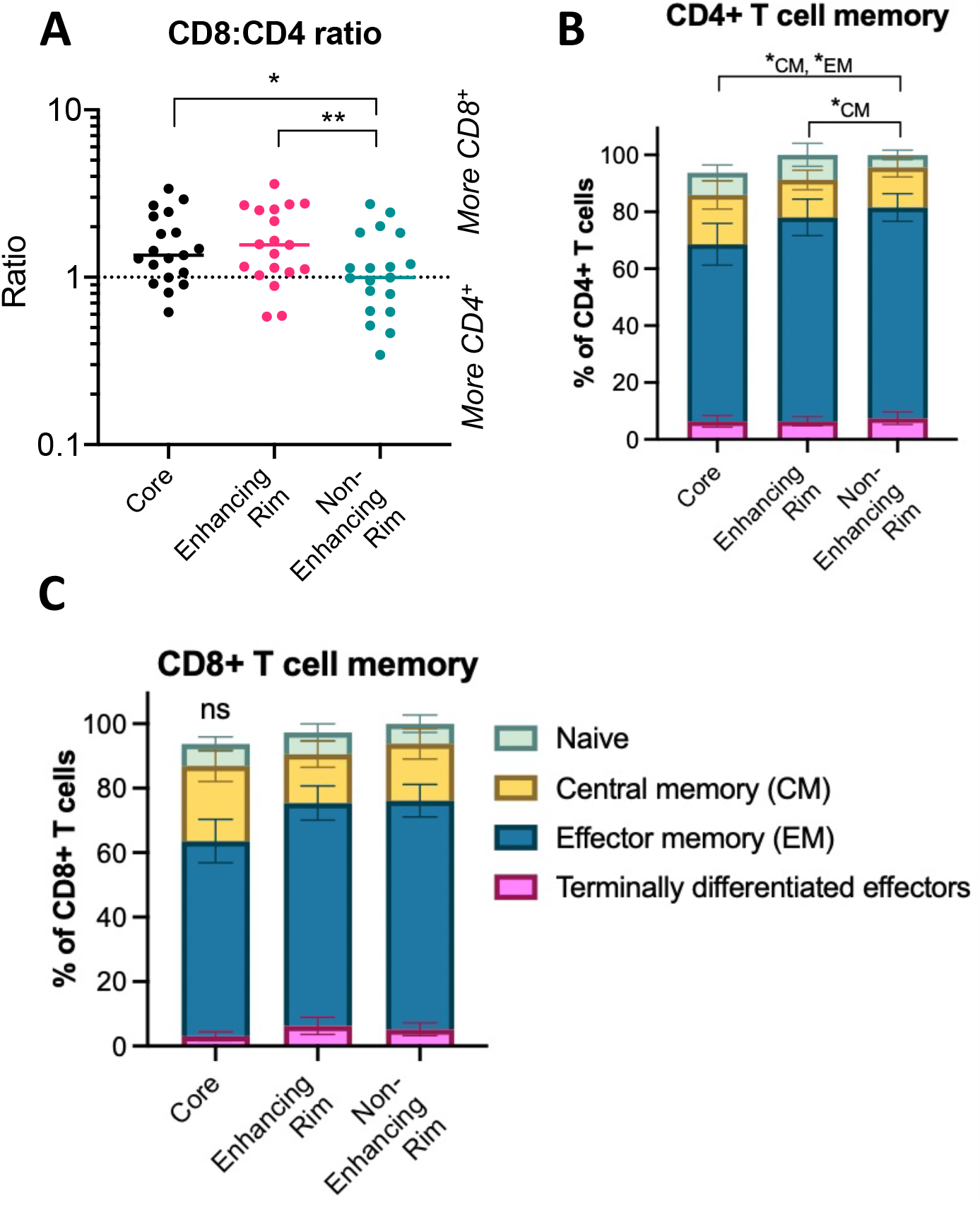
NCE has lower CD8:CD4 T cell ratio, with more abundant CD4+ T effector memory cells. (**A**) CD8:CD4 T cell ratio by flow cytometry. Mixed-effects model ANOVA. (**B**) CD4+ and (**C**) CD8+ T cell memory cell subsets. 2-way ANOVA. *p<0.05; ns – not significant.

**Figure 3:**
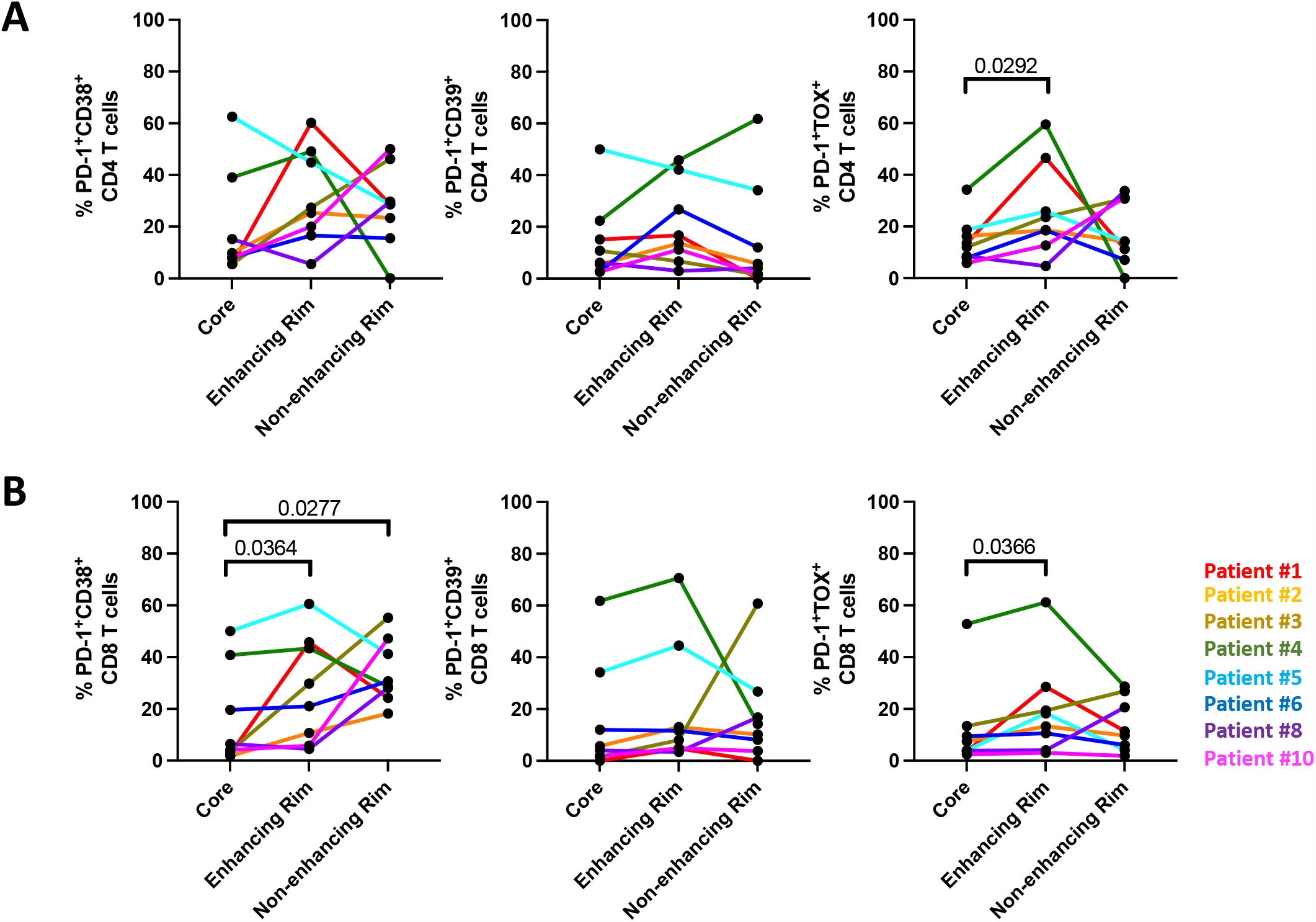
Increased PD-1+TOX+ T cells in CE tumor. (**A**) Abundance of CD38^+^PD-1^+^, CD39^+^PD-1^+^, TOX^+^PD-1^+^ in CD4^+^ T cells in each tumor region, as a percent of total cells. Each line indicates individual patients. Paired t-test. (**B**) Abundance of CD38^+^PD-1^+^, CD39^+^PD-1^+^, TOX^+^PD-1^+^ in CD8^+^ T cells in each tumor region, as a percent of total cells. Each line indicates individual patients. Paired t-test.

**Figure 4:**
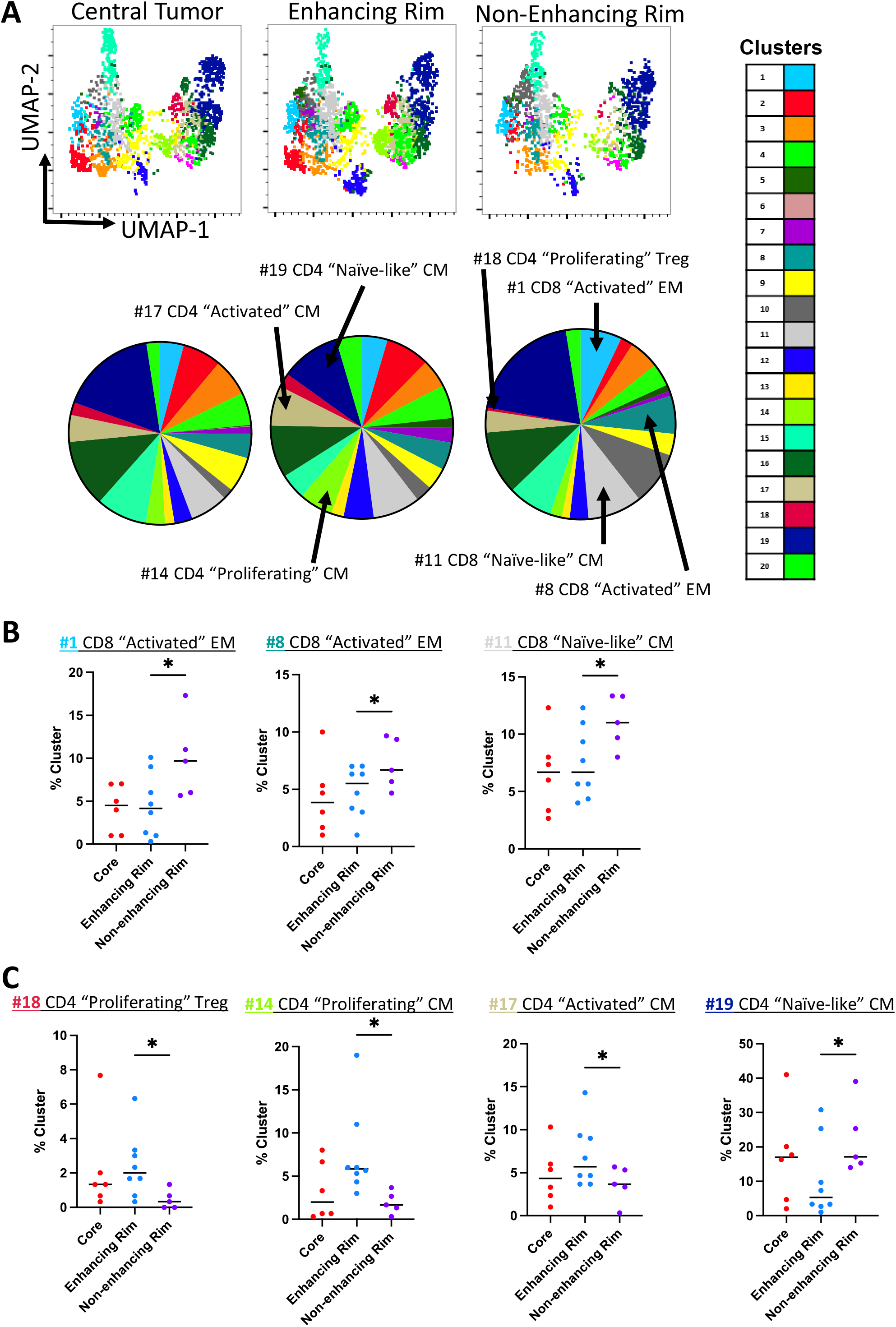
Differential abundance of T cell subsets in NCE revealed by unsupervised clustering analysis. (**A**) Cluster frequency within each region. (**B**) Differentially abundant CD8+ T cell clusters. (**C**) Differentially abundant CD4+ T cell clusters. Paired t-test.

Given that overall T cells found in the NCE regions resembled those found in the CE, we went on to assess their functional capacity in response to *in vitro* nonspecific stimulation with PMA/ionomycin in a subset of patient samples. We found that upon stimulation, both CD8^+^ and non-Treg CD4^+^ (i.e. FoxP3^-^) T cells retained the capacity to produce Granzyme B, TNFα, and IFNγ (**Supplementary Figure S4**).

## DISCUSSION

Findings from this study demonstrate that navigation-assisted sampling and semi-automated tissue collection, stabilization, and preservation using the proposed workflow system is a safe and feasible technique to obtain viable cells for evaluation of immune spatial heterogeneity of GBM. By having a high viability of cells in each specimen, it can increase the volume of usable target tissue collected. Additionally, given the filter design of the tissue acquisition system, minimal specimen (both tumor and immune components) gets discarded through the suction, as can be standard in normal neurosurgical practice. This workflow can be easily standardized across centers and enable safe transport of usable tissue for appropriate analysis regardless of the hospital laboratory capabilities and location, especially for smaller centers.

Our phenotypic and functional analysis of T cells in the NCE suggest that the tumor-associated immune response extends beyond the tumor core and CE, which together comprise the regions of standard-of-care GBM resection. Thus, as an interface of brain and infiltrative tumor, the NCE shows signs of tumor-associated immune alterations, with T cells expressing high levels of activation and exhaustion markers seen in the CE and core regions, despite being fewer in number. There are limited studies published on these findings. It was recently observed using MRI characteristics that the ratio of recruited vs. resident macrophages differed in terms of immunomodulatory activities^9^. This study also found that bone marrow-derived macrophages decrease from the central to the marginal area of the tumors, and microglia cells increase from the necrosis and core to the margin, the former of which is similar to our results. However, this study did not assess lymphocytes in their published results, nor did they assess the functional characteristics of these myeloid cells. Nevertheless, their results suggest that spatial differences in immune infiltrates exist in GBM, and that radiomic profiling using MRI has the potential to non-invasively characterize the heterogeneity of the immunosuppressive microenvironment.

Spatially registered sampling brings about several exciting possibilities with the advent of advanced molecular profiling of GBM tumors and the immune microenvironment, all of which are dependent on accurate analysis of viable cells. GBM tumors have been found to be spatially and temporally heterogenous, creating specific niches with transcriptional adaptations influenced by the tumor microenvironment^10^. As GBM progression typically occurs in a region continuous with the resection cavity, prediction of recurrence-associated genetic and microenvironmental changes and treatments targeted to that specific tumor cell niche may be possible with analysis of spatially-acquired samples. This can be invaluable as individualized immunotherapeutic approaches can be made for each patient, especially with the increasing number of clinical trials utilizing targeted therapies. Additionally, this type of immunosampling may identify potential targetable immune deficits that could be overcome with a specific therapeutic approach (e.g. recurrence region was identified to have exhausted T cells through a specific immune checkpoint receptor). While there is literature supporting supra-total resection (beyond the CE boundary on MRI) to improve outcomes in GBM, boundaries can be limited by critical brain regions that are unable to be removed due to likelihood of permanent post-operative deficit (i.e. eloquent regions), resulting in subtotal resection or only being able to biopsy^7^. Therefore, these results may help inform immunomodulatory strategies needed to target any residual, infiltrating non-enhancing tumor.

### Limitations of the Study

While our study was prospective in nature, limitations of our study include a small number of patients, lack of a normal brain control region, and lack of correlation to outcomes data to identify prognostic factors. A true control region of the brain would ideally require distant or contralateral sampling, which is not feasible due to risk-related ethical reasons. Creation and validation of preclinical animal models could help to inform this point, and design future human studies based off of this work. Further long term follow up and detailed molecular profiling of the tumor specimens will allow us to correlate these findings with patient outcomes.

## Data Availability

All data produced in the present study are available upon reasonable request to the authors

## Acknowledgements

We would like to thank Mary McGraw and Sadie Johnson (Cleveland Clinic) for coordination of tissue procurement. We also thank NICO Corporation for kindly providing the NICO Myriad and Tissue Preservation System, funding for the conduct of the study and for discussions throughout the study.

## Authors’ Contributions

Conceptualization, MMG, DCW, MSA, JDL, AMM; Methodology, MMG, DCW, MSA, JDL, AM, KC; Investigation, MMG, DCW, JL, DB, AM, TA, AMM, KC; Writing – Original Draft, MMG and DCW; Writing – Review & Editing, JL, DB, AL, TA, JM, TC, MSA, JDL, AMM, KC; Funding Acquisition, JDL, MSA, AMM; Resources, JM, JDL, AMM; Supervision, JM, TC, MSA, JDL, AMM

## Inclusion and Diversity Statement

We support inclusive, diverse, and equitable conduct of research.

## MATERIALS AND METHODS

### Clinical study design and objectives

This is a single site, interventional cohort study to assess a standardized method for spatially-registered intraoperative sampling of viable tumor tissue using the NICO Myriad and Tissue Preservation System (TPS) with MRI guidance in 20 prospectively-enrolled GBM patients undergoing surgical resection (NCT04545177). The primary objective of the study was to assess feasibility of the sampling process and viability of each sample’s immune cell population as measured by flow cytometry. The exploratory objectives of the study were to assess the spatial immune heterogeneity of GBM tumor specimens obtained during surgery by flow cytometry in the NCE, CE, and tumor core regions.

### Patient selection

Patients were enrolled if they had a newly-diagnosed or recurrent WHO grade IV, IDH-wildtype glioblastoma (histologically-confirmed via frozen or previous pathology) and were clinically indicated to undergoing resection. All subjects were at least 18 years of age, had a CE tumor volume of at least 15 cc on the preoperative MRI, and a Karnofsky Performance status ≥ 70. One patient was excluded from analysis after enrollment, due to post-operative diagnosis of IDH-mutant tumor.

### MRI-guided tumor segmentation and spatial mapping

Utilizing preoperative stereotactic MRI, the tumor was divided into sectors based on radiographic and/or geographic regions which were accessible/resectable using neuronavigation during surgery. Tumor regions selected for sampling were from 3 separate regions: the necrotic core of tumor, the solid CE rim, and the adjacent NCE, T2/FLAIR abnormal region outside of the enhancing rim **(Figure 1B)**. Priority was given to selecting maximally distinct tissue regions based on geographic location and radiographic findings. Regions were annotated before surgery on the Brainlab Cranial Elements software (Brainlab AG, Munich, Germany).

### Surgical methods

Surgery was performed in the standard fashion with neuronavigation using the Brainlab Cranial software. Annotated regions were localized and the tissue surgically harvested by the neurosurgeon using the navigable NICO Myriad™, an FDA cleared suction/microscissor device that transects tissues and suctions the bio-specimen into a sterile collection chamber, the automated Tissue Preservation System (TPS, NICO Inc., Indianapolis, IN). The closed, sterile, specimen collection chamber is affixed with a 263um filter and housed within a styrofoam chiller. Harvested tissue was held within the collection chamber and perfused with preservation solution (lactated ringers). The chilling apparatus kept bio-specimens at 4-8 □C. Bio-specimen collection chambers were labeled with study-specific identifiers (protocol number, patient ID, date of collection) and unique sector of the tumor, as each sector had its own collection chamber. Labeled samples were then transported in a closed, controlled temperature environment from the operating room to the pathology laboratory for analysis when appropriate.

### Tissue processing

Tumor specimens were kept in the TPS enclosure on ice and transported to the research laboratory within 45 min of acquisition. All subsequent tissue manipulation was carried out in a biosafety cabinet. The enclosures were sequentially opened, and tissue manually collected into a 10cm Petri dish by scraping the collection mesh using flat-end forceps. Half of the tissue was divided into 3-5 mm pieces, flash frozen in liquid nitrogen, and stored at -80°C. The remaining tissue was used to generate single cell suspension. Specifically, tissue was moved Miltenyi gentleMACS C-tubes (up to 1 g per tube) containing 5 mL of digestion solution (Miltenyi human Tumor Dissociation Kit), using the kit components suggested by the manufacturer for downstream flow cytometry. The C-tubes were then connected to the gentleMACS Octo-dissociator with heaters (Miltenyi) and dissociated using the 37_h_TDK_1 device program. Digested material was strained through a 70 um SmartStrainer (Miltenyi) into a 50 mL conical tube. Any residual tissue clumps on the strainer mesh were manually smashed onto the mesh using the back of a 1 mL syringe plunger, and strainer mesh was finally washed with 10-20 mL of RPMI medium. Cell suspension was pelleted by centrifugation at 300 x *g* for 5 min. Cell pellets were resuspended in ∼5 mL of 1X RBC lysis buffer (Biolegend) per 1 g of tissue input, and incubated at room temperature for 5 min, manually gently agitating the cell suspension every 1-2 minutes. The lysis reaction was stopped by addition of 25 mL of PBS, and cells were again pelleted by centrifugation. Cell pellet was resuspended in RPMI and counted with Trypan blue exclusion. Depending on cell yield, 2 to 20 million cells were withdrawn and moved to microtubes for immediate immunophenotyping (*below*). The rest were pelleted by centrifugation and resuspended in serum-free freezing medium (ATCC) for freezing using a rate freezer, and subsequent storage of the frozen single-cell suspensions in liquid nitrogen freezer.

### Flow cytometric analysis

Aliquots of freshly digested single-cell suspensions in microtubes (*see above*) were pelleted by centrifugation at 300 x *g* for 5 min, resuspended in 1 mL of PBS, and pelleted again. Cell pellets were resuspended in 200 μL of Live/Dead Blue solution (1:1,000 in PBS, Invitrogen), and incubated on ice for 10 min, protected from light. 750 μL of FACS buffer (2% bovine serum albumin [BSA, Sigma] in PBS) was added to quench the reaction, and cells were pelleted by centrifugation. Cells were resuspended in 150 μL of human Fc receptor blocking reagent (1:50 in FACS buffer, Miltenyi), and incubated for 15 min on ice, protected from light. Sample divided to 2 (for staining with different antibody cocktails, below), and pelleted by centrifugation. Each cell pellet was then resuspended 100 μL of antibody cocktail prepared in Brilliant Stain Buffer (BD Biosciences) (**Supplementary Table S1**), and incubated for 20 min on ice, protected from light. 500 μL of FACS buffer was added, and cells were pelleted. Cell pellet was resuspended in fixation/permeabilization buffer (FoxP3 Fix/Perm set, Ebioscience), and incubated at 4°C for 30 min or overnight. 1 mL of permeabilization buffer from the same kit, and cells were pelleted by centrifugation at 400 x *g* for 5 min. Cell pellets were resuspended in 100 μL of intracellular antibody cocktails in permeabilization buffer (**Supplementary Table S1**), and incubated for 20 min at room temperature. 1 mL of permeabilization buffer was added, and cells were pelleted by centrifugation at 400 x g for 5 min. Cell pellet was resuspended in FACS buffer and stored at 4°C until flow cytometric analysis on Fortessa LSR II (BD Biosciences) or Cytek Aurora (Cytek Biosciences). Data was analyzed using FlowJo X (BD Biosciences) using gating strategies and unsupervised computational high dimensional analysis outlined in **Supplementary Figures S1-S3**. Biobanked samples were similarly stained upon thawing with T cell focused panels, described in **Tables S2-S3**.

### In vitro T cell functional assay

In vitro T cell functional assay was performed on cryopreserved single-cell suspension. Thawed cell were washed and resuspended in pre-warmed RPMI media supplemented with 10% FBS, 1% penicillin/streptomycin and GlutaMax (Gibco). Cells were stimulated in the presence of stimulation cocktail (PMA/ionomycin, brefeldin A, monensin) (eBioscience) for 5 hours. Then cells were harvested and subjected for flow cytometry analysis as described above.

### Statistics

Statistical analysis was performed on GraphPad Prism 8.4. Comparison of viability and individual cell populations was by paired non-parametric analysis (Friedman Test), with post-hoc Dunn’s multiple comparison test. Comparison of T cell memory phenotypes was by 2-way ANOVA with post-hoc Holm-Sidak test for multiple comparisons. Comparison of clusters from unsupervised analysis was by paired parametric analysis (paired T test).

### Study approval

All patients provided informed consent for study participation prior to enrollment. The clinical trial protocol was reviewed and approved by the Case Comprehensive Cancer Center’s Protocol Review and Monitoring Committee (CASE8319), the Cleveland Clinic Institutional Review Board (#20-499), and is registered on ClinicalTrials.gov (NCT04545177).

