## Supplemental data for "Spatial immunosampling of MRI-defined glioblastoma regions reveals immunologic fingerprint of non-contrast enhancing, infiltrative tumor margins"

**Supplementary Data**

### Table S1: Patient Characteristics.

| ID | Age (decade) | Sex | Laterality | Lobe | CE Volume (Preop  MRI, cm^3^) | IDH Mutated | 1p Del | 19q DeL | MGMT Methylated | Ki_67 (%) | EGFR Amplified | p53 Percent Reactivity |
| --- | --- | --- | --- | --- | --- | --- | --- | --- | --- | --- | --- | --- |
| 1 | 50s | F | L | Temporal | 147.1 | No | No | No | Yes | 50 | No | 80 |
| 2 | 80s | M | R | Occipital | 80.1 | No | Yes | No | Yes | 50 | No | 10 |
| 3 | 70s | M | R | Parietal | 17.5 | No | No | No | No | 40 | Yes | 5 |
| 4 | 60s | M | L | Parietal | 15.6 | No | No | No | No | 20 | No | 75 |
| 5 | 80s | M | R | Occipital | 68.3 | No | No | No | No | 80 | No | 80 |
| 6 | 60s | F | L | Parietal | 32.6 | No | Yes | No | No | 40 | No | 10 |
| 7 | 60s | F | L | Frontal | 20.7 | No | No | No | Yes | 30 | No | 10 |
| 8 | 70s | M | R | Temporal | 29.1 | No | No | No | No | 40 | No | 5 |
| 9 | 80s | M | L | Parietal | 24.6 | No | No | No | No | 40 | No | 10 |
| 10 | 60s | F | R | Parietal | 29.5 | No | No | Yes | No | 25 | No | 20 |
| 11 | 60s | F | L | Temporal | 17.2 | No | No | Yes | No | 25 | No | 50 |
| 12 | 50s | F | L | Occipital | 25.5 | No | Yes | No | No | 25 | Yes | 20 |
| 13 | 50s | F | L | Frontal | 37.7 | No | No | No | No | 80 | Yes | 20 |
| 14 | 60s | F | R | Frontal | 17.5 | No | Yes | No | No | 25 | No | 10 |
| 15 | 60s | F | L | Parietal | 16.2 | No | No | No | No | 70 | No | 20 |
| 16 | 60s | M | L | Temporal | 38.5 | No | No | No | No | 15 | No | 50 |
| 17 | 60s | M | R | Temporal | 18.5 | No | No | No | No | 50 | Yes | 30 |
| 18 | 60s | F | R | Temporal | 36.1 | No | No | No | No | 50 | Yes | 10 |
| 19 | 60s | F | R | Temporal | 66.3 | No | No | No | Yes | 70 | No | 90 |

Abbreviations: CE – contrast-enhancing, IDH – isocitrate dehydrogenase, DEL – deletion, MGMT – O(6)-methylguanine-DNA methyltransferase; EGFR – epidermal growth factor receptor

### Supplementary Figure S1: Gating strategy (fresh single-cell analysis).

**A**

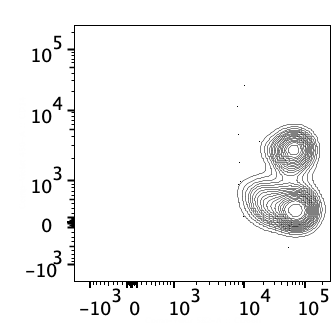

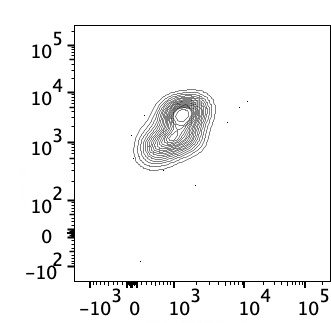

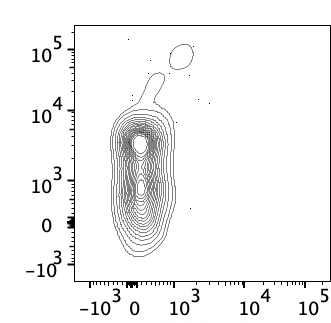

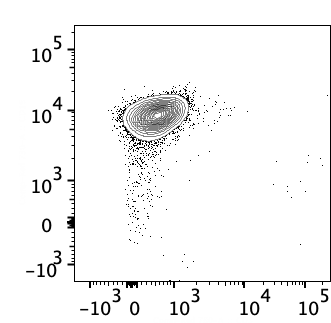

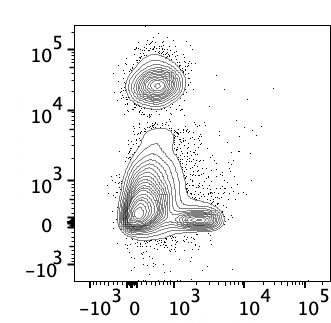

CD66b

CD45

HLA-DR

CD3

CD16

CD3

CD11b

CD33

CD14

CD66b

***Neutrophils***

*CD66b+*

*CD16^low/-^*

*HLA-DR^low/-^*

*CD11b^+^CD33^+^*

***gMDSC***

*CD16^hi^*

*CD14^low/-^*

*CD66b^+^*

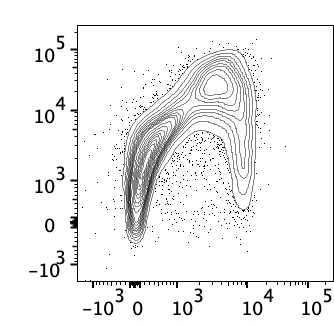

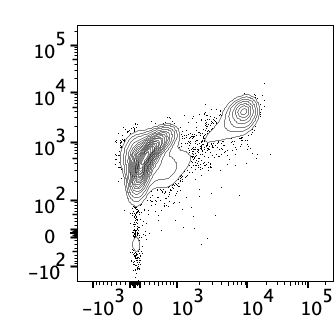

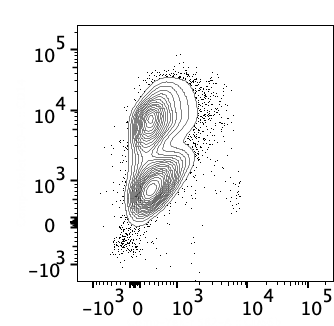

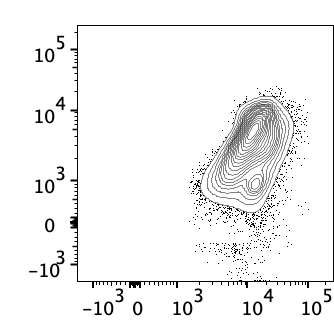

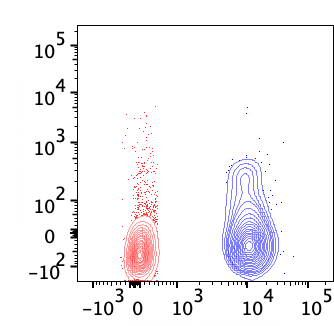

CD11b

CD20

*CD11b^-^*

***CD11b gate reference***

***T cells***

***B cells***

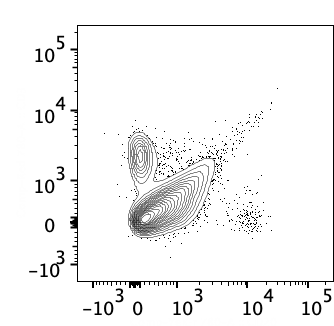

***T cells***

***B cells***

*CD3^+^*

*CD20^+^*

CD3

CD20

*HLA-DR^hi^*

HLA-DR

CD33

CD11b

CD33

*CD11b^+^CD33^+^*

CD14

CD66b

*CD14^hi^CD66b^lo/-^*

***mMDSC***

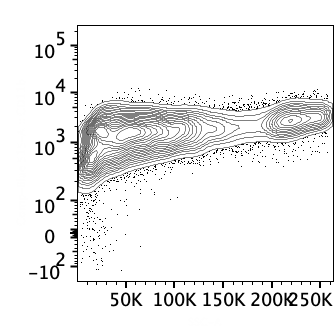

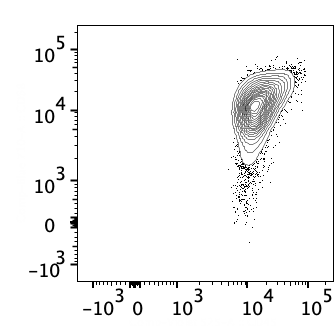

CD11b

SSC

50K

100K

150K

200K

250K

CD33

CD45

CD68

CD45

*CD11b^+^*

*CD45^dim^*

***Microglia***

*CD45^hi^*

*CD68^+^*

***Macrophages***

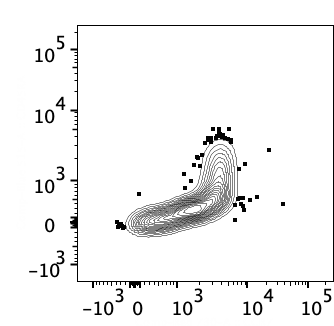

CD45RA

CCR7

***CM***

***EM***

***T_eff_***

***N***

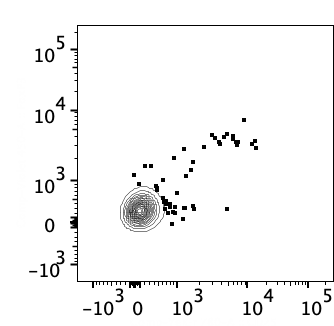

FoxP3

CD25

***Treg***

*FoxP3^+^CD25^+^*

**B**

**C**

**D**

*--Figure S1 caption---*

Events in all panels were already gated for cells (SSC-FSC), viability (Live/Dead staining) and CD45 expression. Contour plot with 5% steps.

(**A**) Granulocyte gating. (**B**) Non-granulocyte gating. Events were already gated for lack of CD66b (see first panel of **Figure S1A**). CD11b expression in T and B cells are shown in upper right, as a reference for subsequent gating. (**C**) T cell memory gating. Events were already gated for CD3, and CD8 expression. Similar gating for CD4+ T cells. Central memory (CM), effector memory (EM), terminally-differentiated T effectors (T_eff_), Naïve (N). (**D**) Treg gating. Events were already gated for CD3, and CD4 expression. T regulatory cells (Treg).

### Supplementary Figure S2: T cell exhaustion marker gating strategy (biobanked samples).

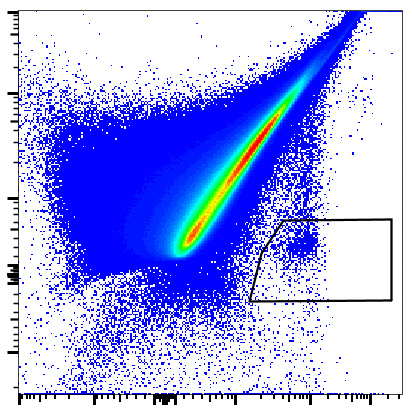

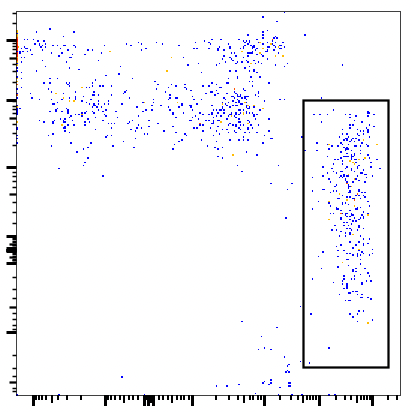

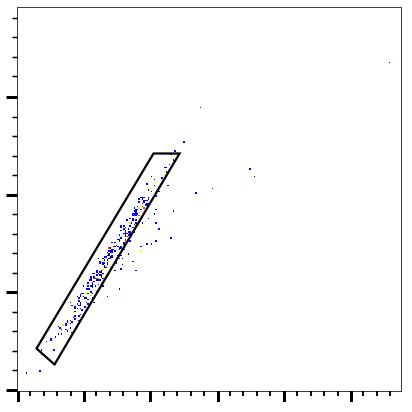

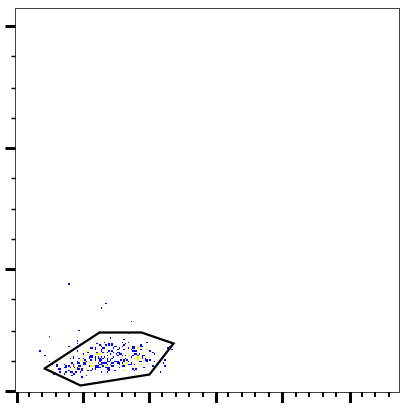

Live/Dead Blue

CD45

Dump

CD3

FSC-Height

FSC-Area

SSC-Area

FSC-Area

**Live Lymphocytes**

**T cells**

**Singlets**

**Intact cells**

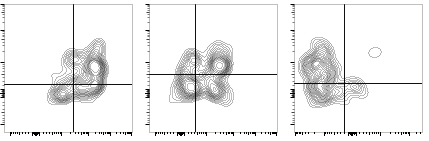

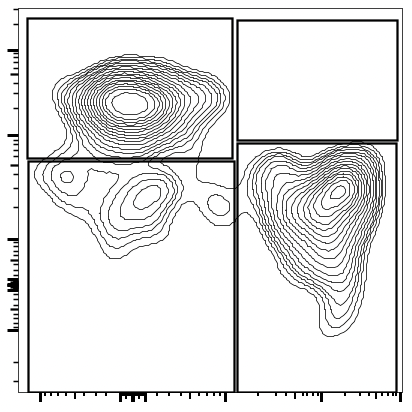

CD4

CD8

**CD8^+^**

**CD8^+^**

**T cells**

**CD4^+^**

**CD4^+^**

**T cells**

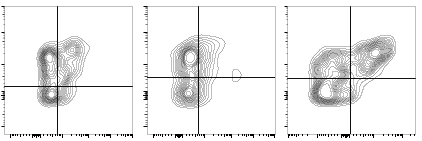

PD-1

CD38

PD-1

CD39

PD-1

TOX

PD-1

CD38

PD-1

CD39

PD-1

TOX

**CD38^+^**

**PD-1^+^**

**CD39^+^**

**PD-1^+^**

**TOX^+^**

**PD-1^+^**

**CD38^+^**

**PD-1^+^**

**CD39^+^**

**PD-1^+^**

**TOX^+^**

**PD-1^+^**

CD19^+^, CD41a^+^, CD14^+^, EpCAM^+^, CD11b^+^ and CD15^+^ cells were excluded from analysis (dump channel). CD4^+^ or CD8^+^ T cells were gated into PD-1^+^CD38^+^, PD-1^+^CD39^+^, PD-1^+^TOX^+^ cell subsets.

### Supplementary Figure S3: Unsupervised clustering of T cells in the core, CE and NCE.

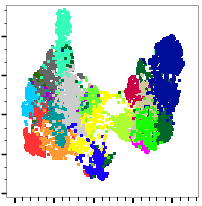

UMAP-1

UMAP-2

**B**

CD4

CD8

CD25

Foxp3

CCR7

CD45RA

CD27

CD28

CD127

CD226

CD69

CD38

ICOS

KI67

HLA-DR

4-1BB

PD-1

TIM3

TIGIT

LAG3

CTLA4

2B4

CD39

TOX

EOMES

TCF1

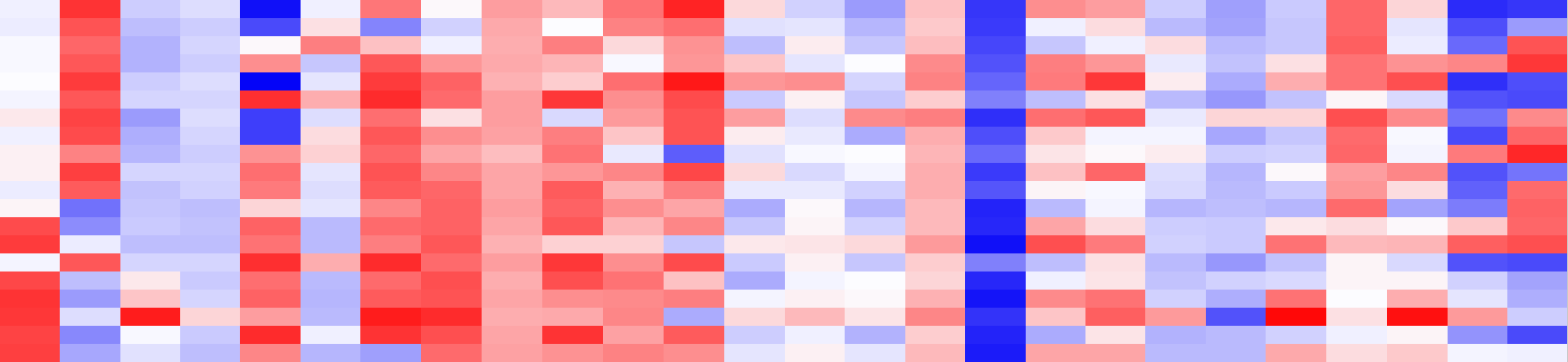

**CD4 “Proliferating” Treg**

**CD4 “Naïve-like” CM (TCF1^hi^)**

CD4 “Effector-like” CM

CD8 Naive-like

CD4 CM

**CD4 “Activated” CM (PD-1^hi^CD69^hi^)**

CD4^-^CD8^-^

CD4 “Activated” CM (EOMES^hi^)

**CD4 “Proliferating” CM (Ki67^hi^)**

CD8 Naïve-like

CD8 “Effector-like” CM

**CD8 “Naive-like” CM (TCF1^hi^)**

CD8 SCM

CD8 ”Proliferating” EM

**CD8 “Activated” EM (CD69^hi^2B4^hi^)**

CD8 Effector

CD8 ”Activated” SCM

CD8 “Activated” EM (Tim3^hi^)

**CD8 “Activated” EM (CD69^hi^PD-1^hi^)**

CD8 Effector

Lineage

Differentiation

Activation

Immune Checkpoints

Effectors

**C**

**A**

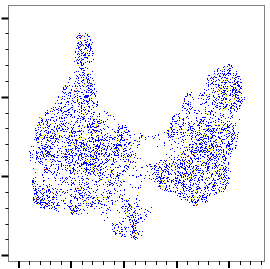

UMAP-1

UMAP-2

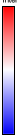

High

Low

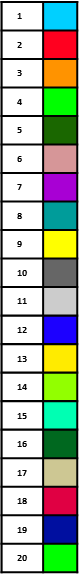

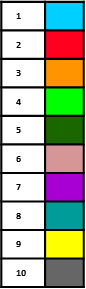

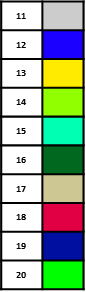

*--Figure S3 caption---*

(**A**) UMAP dimensionality reduction exhaustion/activation T cell panel in subset of 9 patients, each with 3 spatially sampled regions. (**B**) FlowSOM algorithm computed 20 clusters that were superimposed on UMAP and identified by numbers and color as indicated. (**C**) Heap-map represents the marker fluorescence intensity associated to each clusters.

### Supplementary Figure S4: T cells in non-enhancing rim retain cytokine-producing ability found in other regions.

**A**

**B**

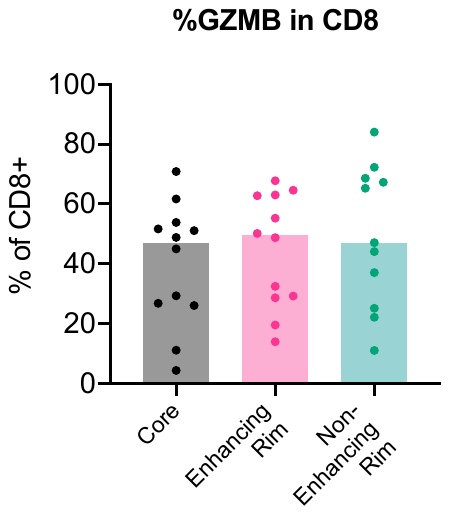

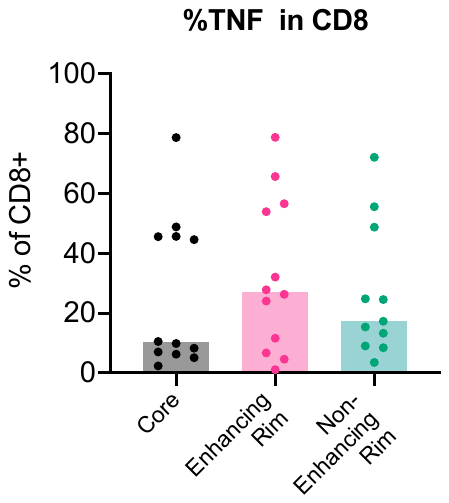

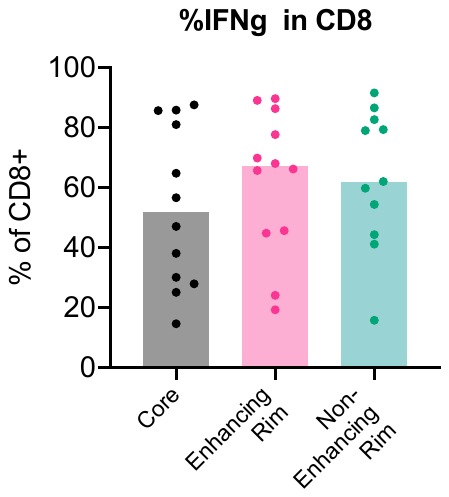

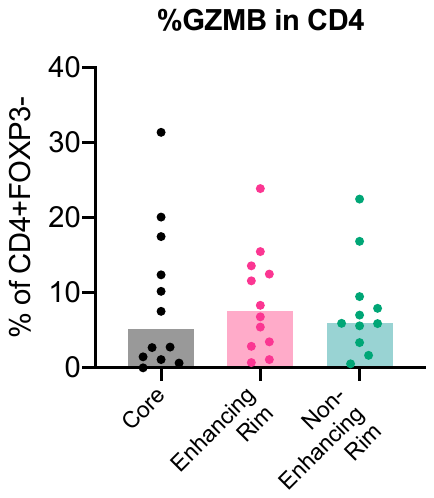

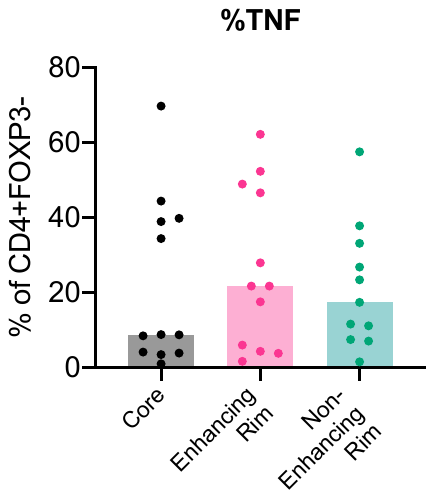

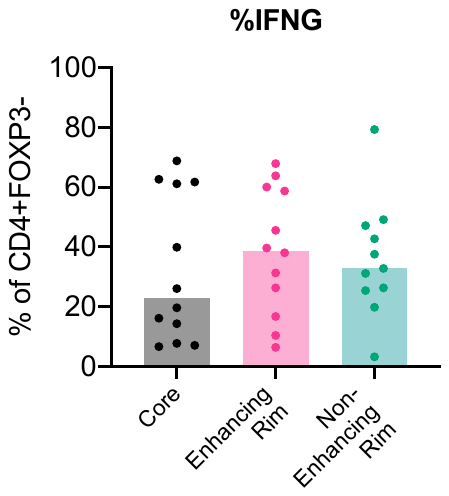

**Granzyme B**

**TNFα**

**IFNγ**

% of CD8^+^

% of CD4^+^FoxP3^-^

Cryopreserved single-cell suspensions from spatially registered tumor samples were stimulated with PMA/ionomycin. Cytokine production was assessed by intracellular staining and flow cytometry, and analyzed separately for (**A**) CD8^+^ and (**B**) Non-Treg CD4^+^ (i.e. FoxP3^-^) T cells.

### Table S2. Flow cytometry panels for immunophenotyping of fresh GBM specimens immediately after tumor digestion.

| **Panel 1** | | | |
| --- | --- | --- | --- |
| *Marker* | *Fluorophore* | *Clone* | *Supplier* |
| **Surface staining** | | | |
| CD11b | FITC | ICRF44 | BD Biosciences |
| CD3 | APC-Cy7 | SP34-2 | BD Biosciences |
| CD33 | PE-CF594 | WM53 | BD Biosciences |
| CD11c | BV650 | 3.9 | Biolegend |
| CD16 | AF700 | 3G8 | BD Biosciences |
| CD20 | PE-Cy7 | 2H7 | BD Biosciences |
| CD14 | Pacific Blue | M5E2 | BD Biosciences |
| CD66b | PE | G10F5 | BD Biosciences |
| Lox-1 | APC | 15C4 | Biolegend |
| CD303 | BV605 | 201A | Biolegend |
| HLA-DR | BV570 | L243 | Biolegend |
| CD45 | V500 | HI30 | BD Biosciences |
| **Intracellular staining** | | | |
| Ki-67 | BV711 | B56 | BD Biosciences |
| CD68 | PerCP-Cy5.5 | Y1/82A | Biolegend |

| **Panel 2** | | | |
| --- | --- | --- | --- |
| *Marker* | *Fluorophore* | *Clone* | *Supplier* |
| **Surface staining** | | | |
| CD45RA | AF488 | HI100 | Biolegend |
| CD3 | APC-Cy7 | SP34-2 | BD Biosciences |
| CD69 | PE-CF594 | FN50 | BD Biosciences |
| PD-1 | BV650 | EH12.1 | BD Biosciences |
| CCR7 | AF700 | G043H7 | Biolegend |
| CD25 | PE-Cy7 | M-A251 | BD Biosciences |
| CTLA4 | APC | BNI3 | BD Biosciences |
| CD4 | BV605 | SK3 | BD Biosciences |
| CD8a | BV570 | RPA-T8 | Biolegend |
| CD45 | V500 | HI30 | BD Biosciences |
| TIGIT | PerCP-Cy5.5 | A15153G | Biolegend |
| **Intracellular staining** | | | |
| Ki-67 | BV711 | B56 | BD Biosciences |
| Eomes | PE | WD1928 | ThermoFisher |
| FoxP3 | V450 | 236A/E7 | BD Biosciences |

### Table S3. Flow cytometry panel for analysis of in vitro T cell functional assays

| **Panel** | | | |
| --- | --- | --- | --- |
| *Marker* | *Fluorophore* | *Clone* | *Supplier* |
| **Surface staining** | | | |
| CD45RA | AF488 | HI100 | Biolegend |
| CD3 | APC-Cy7 | SP34-2 | BD Biosciences |
| CXCR5 | BUV563 | RF8B2 | BD Biosciences |
| PD-1 | BV650 | EH12.1 | BD Biosciences |
| CCR7 | AF700 | G043H7 | Biolegend |
| TIM3 | BV510 | F38-2E2 | Biolegend |
| CTLA4 | APC | BNI3 | BD Biosciences |
| CD4 | BUV737 | SK3 | BD Biosciences |
| CD8a | BV570 | RPA-T8 | Biolegend |
| CD45 | BUV395 | HI30 | BD Biosciences |
| TIGIT | PerCP-Cy5.5 | A15153G | Biolegend |
| CD39 | SB702 | A1 | invitrogen |
| KLRG1 | PE-Cy7 | 2F1/KLRG1 | Biolegend |
| **Intracellular staining** | | | |
| FoxP3 | V450 | 236A/E7 | BD Biosciences |
| TNFa | BV421 | MAb11 | Biolegend |
| Tbet | BV605 | 4B10 | Biolegend |
| Granzyme B | PE-CF594 | GB11 | BD Biosciences |
| TOX | PE | TXRX10 | invitrogen |
| TCF1 | AF647 | C63D9 | Cell Signaling Technology |
| IFNg | BV786 | 4S.B3 | BD Biosciences |

### Table S4. Flow cytometry panel for analysis of T cell exhaustion/activation (biobanked samples).

| **Panel** | | | |
| --- | --- | --- | --- |
| *Marker* | *Fluorophore* | *Clone* | *Supplier* |
| CD28 | BUV395 | CD28.2 | BD |
| Live/Dead | Live/Dead Blue | Fixable Viability Stain 510 | BD |
| CXCR5 | BUV563 | RF8B2 | BD |
| CD25 | BUV615 | 2A3 | BD |
| CD226 (DNAM-1) | BUV661 | DX11 | BD |
| PD-1 | BUV737 | EH12.1 | BD |
| CD3 | BUV805 | HIT3a | BD |
| ICOS | efluor450 | ISA-3 | ThermoFisher |
| CD127 | BV480 | HIL-7R-M21 | BD |
| CD19 | BV510 | SJ25C1 | BD |
| CD41a | BV510 | HIP8 | BD |
| CD14 | BV510 | M5E2 | BD |
| EpCAM (CD326) | BV510 | EBA-1 | BD |
| CD11b | BV510 | D12 | BD |
| CD15 | BV510 | W6D3 | BD |
| HLA-DR | Spark Violet 538 | L243 | Biolegend |
| CD45RA | BV570 | HI100 | Biolegend |
| CD244 (2B4) | BV605 | C1.7 | Biolegend |
| T-bet | BV650 | O4-46 | BD |
| Tim3 | BV711 | 7D3 | BD |
| CD39 | BV750 | TU66 | BD |
| CD4 | qdot800 | S3.5 | ThermoFisher |
| CD38 | BB515 | HIT2 | BD |
| CD8 | Spark blue 550 | SK1 | Biolegend |
| CD45 | AF532 | HI30 | ThermoFisher |
| FoxP3 | BB700 | 236A/E7 | BD |
| Tigit | PerCP Efluor710 | MBSA43 | ThermoFisher |
| CCR7 | Spark yg 581 | G043H7 | Biolegend |
| CD69 | AF561 | FN50 | ThermoFisher |
| TOX | PE | Rea473 | Miltenyi |
| Eomes | PE-eFluor610 | WD1928 | ThermoFisher |
| CTLA4 | PE-Cy5 | BNI3 | BD |
| KLRG1 | PE-Cy7 | 2F1/KLRG1 | Biolegend |
| TCF1 | AF647 | C63D9 | Cell Signaling |
| LAG3 | AF660 | 3DS223H | ThermoFisher |
| KI67 | AF700 | B56 | BD |
| 4-1BB | APC efluor 780 | 4B4 (4B4-1) | FisherScientific |
| CD27 | APC Fire 810 | QA17A18 | Biolegend |
